# Antipsychotic prescribing and mortality in people with dementia before and during the COVID-19 pandemic: retrospective cohort study

**DOI:** 10.1101/2023.02.18.23286127

**Authors:** Christian Schnier, Aoife McCarthy, Daniel R Morales, Ashley Akbari, Reecha Sofat, Caroline Dale, Rohan Takhar, Mamas A. Mamas, Kamlesh Khunti, Francesco Zaccardi, Cathie LM Sudlow, Tim Wilkinson, the CVD-COVID-UK/COVID-IMPACT Consortium

## Abstract

**Background:** Antipsychotic drugs have been associated with increased mortality, stroke and myocardial infarction in people with dementia. Concerns have been raised that antipsychotic prescribing may have increased during the COVID-19 pandemic due to social restrictions imposed to limit the spread of the virus. We used multisource, routinely-collected healthcare data from Wales, UK, to investigate prescribing and mortality trends in people with dementia before and during the COVID-19 pandemic.

**Methods:** We used individual-level, anonymised, population-scale linked health data to identify adults aged ≥60 years with a diagnosis of dementia in Wales, UK. We explored antipsychotic prescribing trends over 67 months between 1^st^ January 2016 and 1^st^ August 2021, overall and stratified by age and dementia subtype. We used time series analyses to examine all-cause, myocardial infarction (MI) and stroke mortality over the study period and identified the leading causes of death in people with dementia.

**Findings:** Of 57,396 people with dementia, 11,929 (21%) were prescribed an antipsychotic at any point during follow-up. Accounting for seasonality, antipsychotic prescribing increased during the second half of 2019 and throughout 2020. However, the absolute difference in prescribing rates was small, ranging from 1253 to 1305 per 10,000 person-months. Prescribing in the 60-64 age group and those with Alzheimer’s disease increased throughout the 5-year period. All-cause and stroke mortality increased in the second half of 2019 and throughout 2020 but MI mortality declined. From January 2020, COVID-19 was the second commonest underlying cause of death in people with dementia.

**Interpretation:** During the COVID-19 pandemic there was a small increase in antipsychotic prescribing in people with dementia. The long-term increase in antipsychotic prescribing in younger people and in those with Alzheimer’s disease warrants further investigation.

**Funding:** British Heart Foundation (BHF) (SP/19/3/34678) via the BHF Data Science Centre led by HDR UK, and the Scottish Neurological Research Fund.

**Research in Context:** *Evidence before this study:* We searched Ovid MEDLINE for studies describing antipsychotic prescribing trends in people with dementia during the COVID-19 pandemic, published between 1st January 2020 and 22nd March 2022. The following search terms were used: (exp Antipsychotic Agents/ OR antipsychotic.mp OR neuroleptic.mp OR risperidone.mp OR exp Risperidone/ OR quetiapine.mp OR exp Quetiapine Fumarate/ OR olanzapine.mp OR exp Olanzapine/ OR exp Psychotropic Drugs/ or psychotropic.mp) AND (exp Dementia/ OR exp Alzheimer Disease/ or alzheimer.mp) AND (prescri*.mp OR exp Prescriptions/ OR exp Electronic Prescribing/ OR trend*.mp OR time series.mp). The search identified 128 published studies, of which three were eligible for inclusion. Two studies, based on data from England and the USA, compared antipsychotic prescribing in people with dementia before and during the COVID-19 pandemic. Both reported an increase in the proportion of patients prescribed an antipsychotic after the onset of the pandemic. A third study, based in the Netherlands, reported antipsychotic prescription trends in nursing home residents with dementia during the first four months of the pandemic, comparing prescribing rates to the timings of lifting of social restrictions, showing that antipsychotic prescribing rates remained constant throughout this period.

*Added value of this study:* We conducted age-standardised time series analyses using comprehensive, linked, anonymised, individual-level routinely-collected, population-scale health data for the population of Wales, UK. By accounting for seasonal variations in prescribing and mortality, we demonstrated that the absolute increase in antipsychotic prescribing in people with dementia of any cause during the COVID-19 pandemic was small. In contrast, antipsychotic prescribing in the youngest age group (60-64 years) and in people with a subtype diagnosis of Alzheimer’s disease increased throughout the five-year study period. Accounting for seasonal variation, all-cause mortality rates in people with dementia began to increase in late 2019 and increased sharply during the first few months of the pandemic. COVID-19 became the leading non-dementia cause of death in people with dementia from 2020 to 2021. Stroke mortality increased during the pandemic, following a similar pattern to that of all-cause mortality, whereas myocardial infarction rates decreased.

*Implications of all the available evidence:* During COVID-19 we observed a large increase in all-cause and stroke mortality in people with dementia. When seasonal variations are accounted for, antipsychotic prescribing rates in all-cause dementia increased by a small amount before and during the pandemic in the UK. The increased prescribing rates in younger age groups and in people with Alzheimer’s disease warrants further investigation.

## INTRODUCTION

There are an estimated 57 million people living with dementia worldwide.^1^ The COVID-19 pandemic has disproportionately affected people with dementia, as they are more likely to contract the infection and develop severe disease compared to people without dementia.^2,3^ Furthermore, social restrictions imposed to limit the spread of the virus may have been associated with worsened neuropsychiatric symptoms in people with dementia.^4^

Antipsychotic medications can be used to treat symptoms of agitation, aggression, distress, and psychosis in people with dementia. However, antipsychotic use has been associated with an increased risk of adverse outcomes, including stroke,^5,6^ myocardial infarction,^7,8^ and death.^9,10^ As a result, many countries have restrictions on the use of these drugs in people with dementia.^11,12^ The UK National Institute for health and Care Excellence (NICE) clinical guidance states that antipsychotic medications should only be used when alternative approaches have failed, and if the patient is in severe distress or is at risk of harming themselves or others.^13,14^

Concerns have been raised that rates of antipsychotic prescribing for people with dementia may have increased during the early months of the COVID-19 pandemic.^15,16^ The National Health Service (NHS) England produces monthly reports on the proportion of people with dementia who are prescribed an antipsychotic drug: this proportion remained constant throughout 2018-2019 but appeared to increase in March 2020, around the time when social restrictions were introduced in England to reduce the spread of COVID-19.^15^ Whilst these group-level data are useful to understand overall trends, they do not provide detailed information on the demographic details, seasonal variation and linked health outcomes associated with medication use in this population. In this study, we therefore used routinely-collected, linked health data from Wales, UK, to explore trends in antipsychotic prescribing and all-cause, myocardial infarction and stroke mortality in people with dementia before and during the COVID-19 pandemic.

## METHODS

### Study design

We conducted a retrospective cohort study using routinely-collected health data. We pre-published the protocol with disease phenotyping codes (https://github.com/BHFDSC/CCU016_01).

### Study period

We defined the study period from 1^st^ January 2016 until 1^st^ August 2021 (67 months). Follow-up for each participant started at the earliest date of dementia diagnosis or, if a participant received a dementia diagnosis prior to the study start date, on 1^st^ January 2016. Follow-up for each participant ended at the earliest of the date of death, deregistration with a SAIL GP or 1^st^ August 2021. For reference, the first COVID-19 case in the UK was in January 2020, and the UK’s first nationwide lockdown to mitigate the spread of the virus was in March 2020.

### Data sources

The CVD-COVID-UK is a UK-wide initiative established to accelerate research on COVID-19 and cardiovascular disease, by facilitating access to linked routinely-collected electronic health record (EHR) data from England, Scotland and Wales.^17^ For this study, we used the CVD-COVID-UK initiative to access Welsh routinely-collected EHR data from the Secure Anonymised Information Linkage (SAIL) Databank (https://saildatabank.com). SAIL contains anonymised, individual-level, population-scale linked routinely-collected EHR and social care data sources for the population of Wales, UK.^18–20^ As one of four nations in the UK, Wales’ population of approximately three million people receive the majority of their healthcare through the NHS. Primary care data are available in SAIL for approximately 86% of the Welsh population. Data are accessed via a remotely-accessible, privacy-protecting trusted research environment (TRE).

We created a population-based cohort of people with dementia using linked, individual-level anonymised data sources, including: primary care general practice (Welsh Longitudinal General Practice [WLGP]), secondary care hospital admissions (Patient Episode Database for Wales [PEDW]), mortality from a combination of Annual District Death Extract (ADDE), Annual District Death Daily (ADDD) and Consolidated Death Data Source (CDDS), Lower-layer Super Output Area (LSOA) of residence and associated Welsh Index of Multiple Deprivation (WIMD) (Welsh Demographic Service Dataset [WDSD]) and care home (CARE) data sources.

### Eligibility criteria

We included patients who were alive and registered with a SAIL general practice (GP) on 1^st^ January 2016 and who received a dementia diagnosis prior to (using all available previous data) or during the study period. We excluded people with a dementia diagnosis before the age of 60, due to the low rates of dementia and mortality in these ages and the low accuracy of dementia diagnostic coding in younger ages.^21,22^.

We used a validated list of codes in primary care (Read version 2) or hospital admissions (International Classification of Diseases version 10 [ICD-10]) data to identify people with dementia (all-cause dementia) (Supplementary Material, Appendix A).^23^ We defined dementia as the presence of ≥1 dementia codes in either data source, using the date of the first dementia code as the date of diagnosis. We did not use a prescription of a dementia medication (donepezil, rivastigmine, galantamine, memantine) to identify dementia cases but, for people with a prescription for a dementia medication prior to the date of their first dementia diagnostic code, we used the date of the first prescription as the date of dementia diagnosis. We defined dementia subtypes (Alzheimer’s disease, vascular dementia, frontotemporal dementia or dementia with Lewy Bodies) as the presence of one or more subtype codes at any point (Supplementary Material, Appendix A). Dementia subtype categories were not mutually exclusive.

### Exposure to psychotropic medications

We defined exposure to antipsychotic medications during the study window using the prescription data, which are part of the Read-coded primary care dataset (Supplementary Material, Appendix B). As a comparison, we also identified exposure to benzodiazepines, an alternative class of drugs that can treat agitation or distress in people with dementia. We included benzodiazepines prescribing to investigate whether any change seen in antipsychotic prescribing was specific to that medication class, or reflected changes in prescribing rates across all psychotropic medications.

### Frailty

We estimated frailty in participants with a minimum of five years of available primary care data prior to the study start date, using the electronic Frailty Index (eFI).^24^ The eFI algorithm uses primary care data to estimate an individual participant’s frailty, defined as ‘fit’, ‘mild’, ‘moderate’ and ‘severe’. We used the preceding five years of primary care data to calculate the eFI.

### Outcomes

We used mortality data to describe trends in all-cause mortality, mortality due to myocardial infarction and ischaemic stroke over the study period. We used fatal ischaemic stroke and myocardial infarction to investigate cerebrovascular and cardiovascular outcomes respectively, to avoid the uncertainty of whether repeated non-fatal stroke or myocardial infarction codes, such as those identified in hospital admissions data, represent recurrent events or the same event coded multiple times. We used the code lists for stroke and myocardial infarction that were developed during the creation of the SAIL Dementia electronic Cohort.^20^

To determine causes of death, we grouped ICD-10 codes using the same method employed by the Office for National Statistics (ONS) when collating UK mortality figures, with the addition of the new COVID-19 ICD-10 codes (U07.1 and U07.2).^25^

### Statistical analysis

We described the demographics of the study cohort and of the subgroup of participants prescribed an antipsychotic drug.

We calculated the age-standardised monthly rate of antipsychotic and benzodiazepine prescribing between 2016-2021 (67 months) in people with dementia. Using time-series analyses the raw data was decomposed into seasonal, trend and random/residual components using a moving average with symmetric window and an additive model. We standardised ages based on the age distribution of the study cohort in January 2020 (the month in which the first COVID-19 case was subsequently identified in the UK), to account for any changes in the age distribution of the cohort over the study period. We also stratified by sex, age (5-year bands), dementia subtype and care home versus community residence. We described the prescribing patterns for all antipsychotic drugs combined, as well as the top three most commonly prescribed antipsychotic drugs (risperidone, olanzapine and quetiapine).

We conducted an age-standardised time series analysis to investigate changes in all-cause mortality, fatal stroke and fatal myocardial infarction between 2016 to 2021. We compared mortality rates in which stroke or myocardial infarction are listed at the primary position on the death certificate, to rates of these diagnoses being listed anywhere on the death certificate. As with antipsychotic prescribing trends, we also stratified by sex, age (5-year bands), dementia subtype and care home residency.

We counted the underlying causes of death in people with dementia who died between 1^st^ January 2020 and 1^st^ August 2021 (end of follow-up). We compared the number of participants for whom COVID-19 was listed as the underlying cause of death to the number in whom COVID-19 was listed anywhere on the death certificate (underlying and secondary).

We used SQL for data management and R (https://www.r-project.org/) for statistical analysis. The statistical code is publicly available on GitHub (https://github.com/BHFDSC/CCU016_01).

## RESULTS

### Study population

We included 57,396 people with dementia in the study (Figure 1), who contributed 101,428 person-years of follow-up. The median observation time per person was 32 months (IQR 12-66). Participant demographics are displayed in Table 1.

**Table 1.** Participant characteristics. Demographics of the whole cohort and subgroup who were prescribed an antipsychotic drug at any
point during follow-up. IQR – interquartile range, eFI – electronic Frailty Index. *Prior to or at the study start date (01/01/2016). †Categories not mutually exclusive, apart from ‘unspecified’ category, which reflects the number of participants with no subtype code. ‡eFI could only be calculated for participants with five years of available primary care data, prior to the study start date.

|  | Number (%) |  |
| --- | --- | --- |
|  | Total cohort | Subpopulation exposed to antipsychotic drugs |
| Total population | 57,396 | 11,929 |
| Total observation time (person-years) | 101,428 | 22,699 |
| Age at study start date, median [IQR] | 82 [76-87] | 80 [64-86] |
| Sex |  |  |
| Female | 35,148 (61) | 7,115 (60) |
| Male | 22,248 (39) | 4,814 (40) |
| Deprivation (quintiles)* |  |  |
| 1 (most deprived) | 10,624 (19) | 2,375 (20) |
| 2 | 11,656 (20) | 2,513 (21) |
| 3 | 11,091 (19) | 2,200 (18) |
| 4 | 11,425 (22) | 2,313 (19) |
| 5 (least deprived) | 12,600 (22) | 2,528 (21) |
| Resident in care home* | 8,271 (14) | 2,206 (19) |
| Dementia subtype† |  |  |
| Alzheimer's disease | 25,407 (44) | 5,738 (48) |
| Vascular dementia | 17,720 (31) | 4,302 (36) |
| Frontotemporal dementia | 571 (1) | 183 (2) |
| Dementia with Lewy Bodies | 1,044 (2) | 388 (3) |
| Unspecified | 17,506 (31) | 2,835 (24) |
| Date of dementia diagnosis |  |  |
| Before 1/1/2016 | 35,940 (63) | 6,580 (55) |
| On or after 1/1/2016 | 21,456 (37) | 5,349 (45) |
| Frailty [eFI]* |  |  |
| Fit | 12,255 (21) | 1,771 (15) |
| Mild | 11,857 (21) | 2,017 (17) |
| Moderate | 5,240 (9) | 956 (8) |
| Severe | 576 (1) | 100 (1) |
| Missing ‡ | 27,468 (48) | 7,085 (59) |
| Previous stroke* | 7,213 (13) | 1,441 (12) |
| Previous myocardial infarction* | 6,267 (11) | 1,242 (10) |

**Figure 1:**
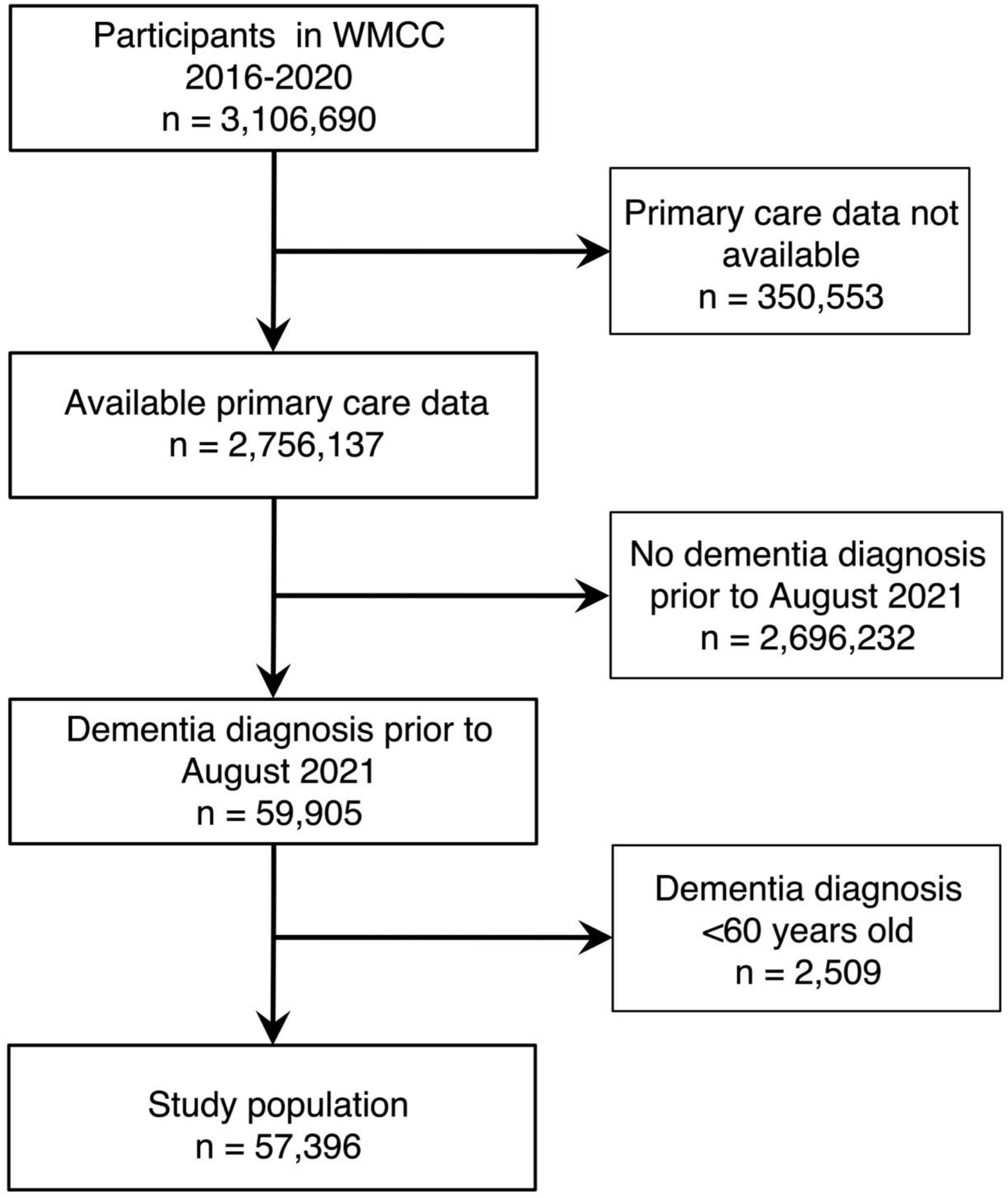
Participant flow diagram.

### Antipsychotic prescribing

Of the 57,396 participants, 11,929 (21%) were prescribed an antipsychotic drug at any point during follow-up. Compared to the total cohort, patients who received an antipsychotic drug were more likely to be younger (median age 80 vs 82 years old; Table 1) and a care home resident at the study start date (19% versus 14%).

Figure 2 shows the age-standardised prescribing rates for all antipsychotic medications and the three most commonly prescribed antipsychotic medications in the population – risperidone, olanzapine and quetiapine. Benzodiazepine prescribing rates are also included by way of a comparison of prescribing trends. Risperidone and olanzapine prescribing rates increased steadily between 2016 and 2021, whereas quetiapine prescribing reduced over the same period (Supplementary Material, Appendix D). Age-stratified antipsychotic prescribing rates appeared relatively static over the entire study period, except for the youngest age group (60-64 years), in whom prescribing rates doubled over the five years (Supplementary Material, Appendix E).

**Figure 2.**
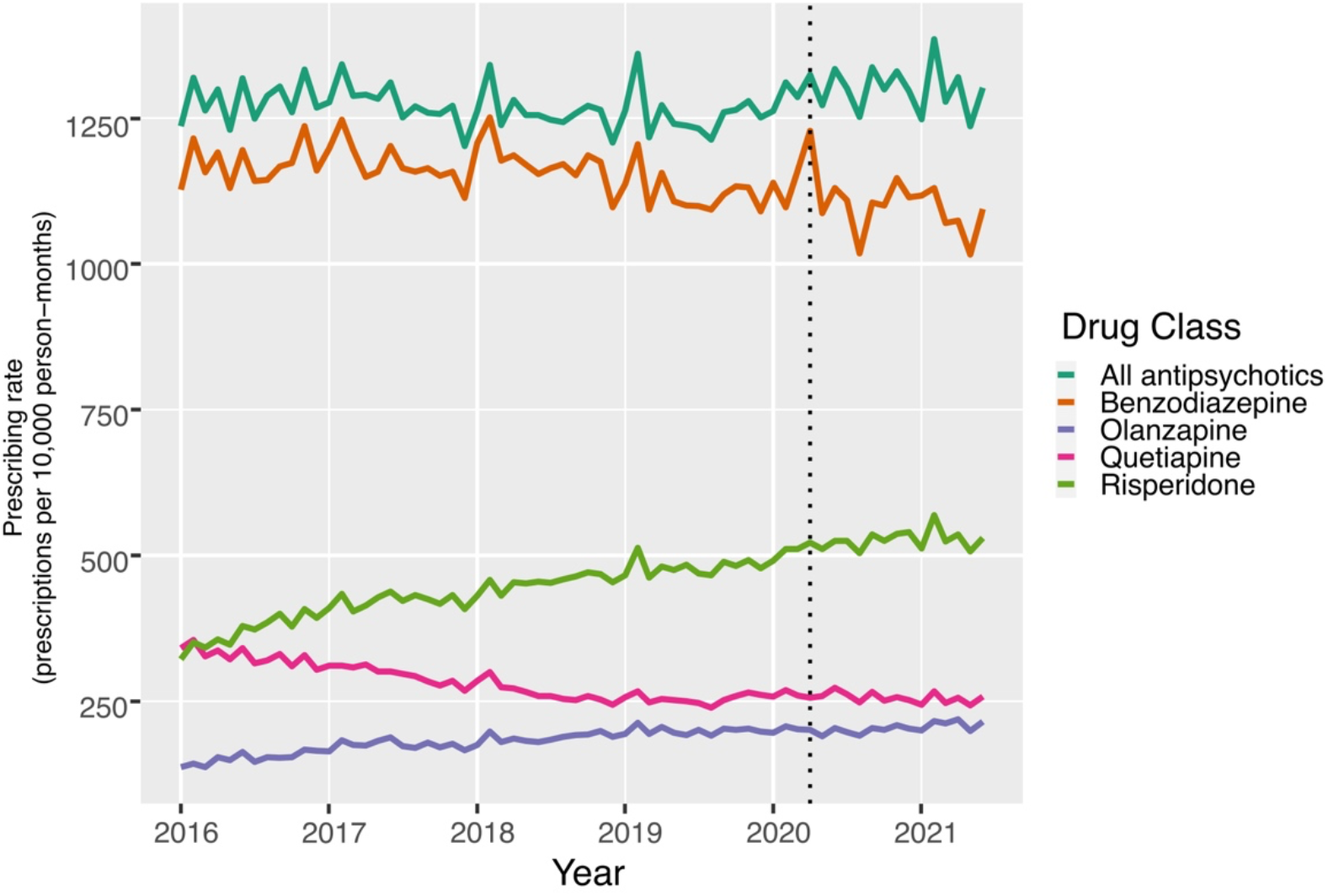
Antipsychotic prescribing rates. Rates are age-standardised. Benzodiazepine prescribing included by way of comparison. Dotted vertical line indicates March 2020, the month of the first UK-wide lockdown to reduce the spread of COVID-19.

In March 2020 (early in the COVID-19 pandemic), 12·9% of people with dementia were prescribed an antipsychotic drug, compared to 12·5% in March 2019. Accounting for seasonal changes, Figure 3 shows a downward trend in antipsychotic prescribing in people with all-cause dementia between 2017 and 2019. Prescribing rates then started to increase in the second half of 2019, continued to increase throughout 2020, and peaked in the second half of 2020. However, the absolute change in prescribing between the lowest and highest rates was relatively small (1253 per 10,000 person-months in March 2019 compared to 1305 per 10,000 person-months in September 2020). Antipsychotic prescribing trends in the subgroup with vascular dementia showed a similar pattern to that of all-cause dementia. In contrast, antipsychotic prescribing in participants with Alzheimer’s disease increased steadily throughout the entire study period, with a steep increase during the second half of 2019, peaking in mid-2020 (Figure 3).

**Figure 3.**
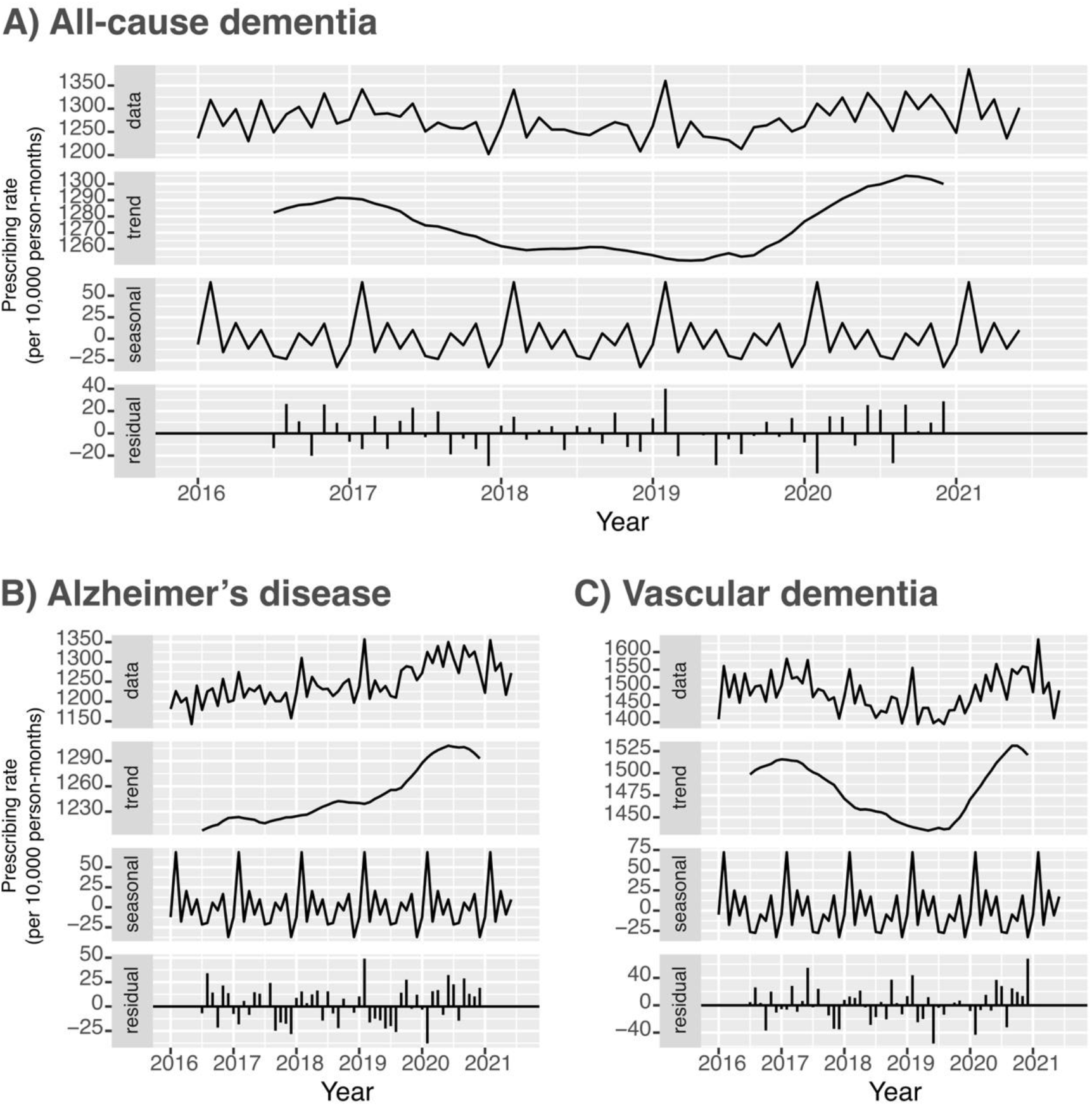
Trends in antipsychotic drug prescribing rates in people with (A) all-cause dementia, (B) Alzheimer’s disease, (C) vascular dementia. Age-standardised, additive time series, accounting for seasonal trends in prescribing. Data – age-standardised prescribing rates. Trend – trend of prescribing rates, accounting for seasonal variations. Seasonal – seasonal trends in prescribing rates during the study period. Residual – difference between the raw data and the seasonal trend.

## Mortality

### All-cause, cardiovascular disease and stroke mortality

During the five-year study period, 35,565 (62%) participants died. The median age of death was 87 years (IQR 82-91). Age-standardised all-cause mortality declined from 2018 until late 2019, then increased sharply and continued to increase throughout early and mid-2020 (Figure 4).

**Figure 4.**
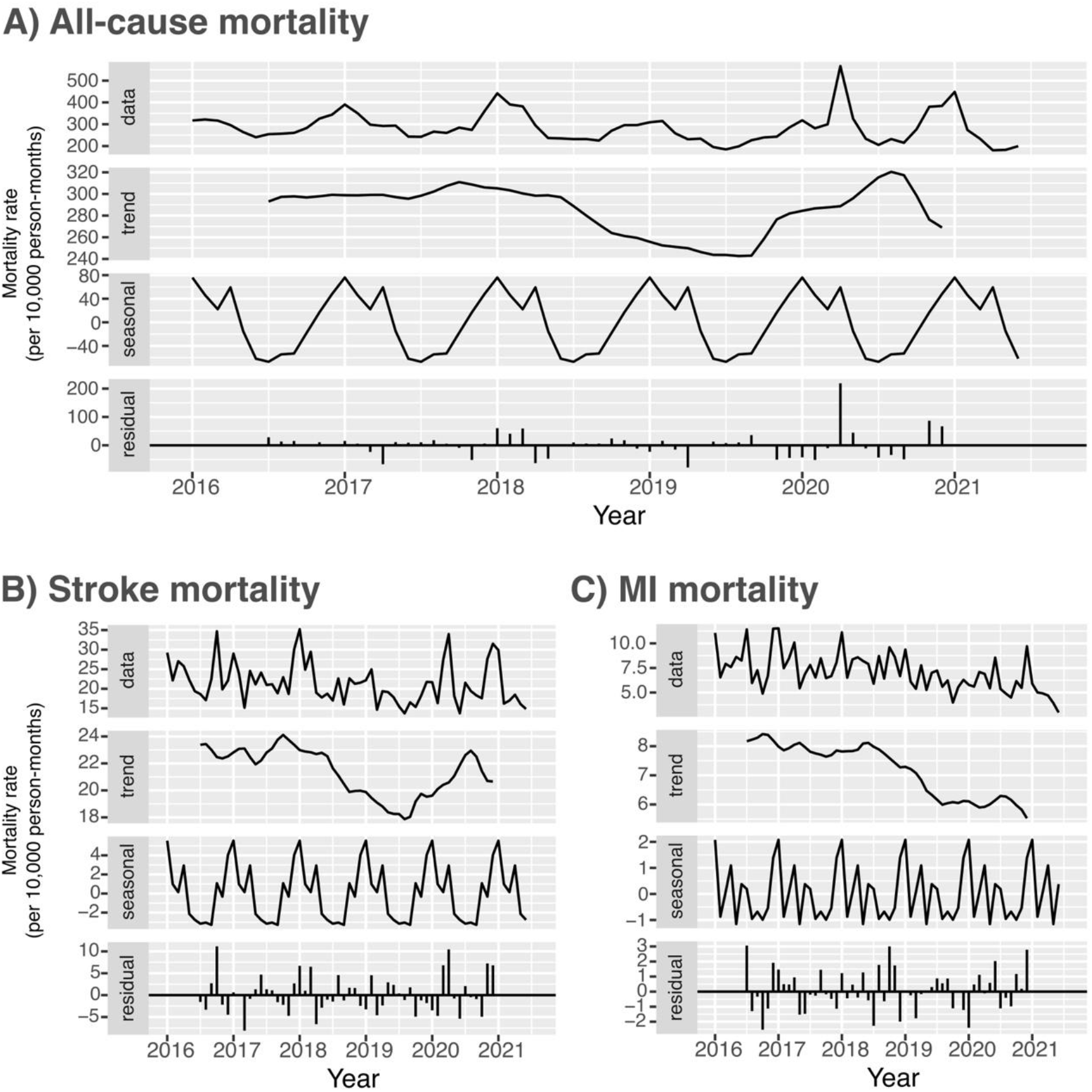
Trends in all-cause and cause-specific mortality rates. Age-standardised, additive time series, accounting for seasonal trends in mortality. Data – age-standardised mortality rates. Trend – trend if mortality rates, accounting for seasonal trends. Seasonal – seasonal trends in prescribing rates during the study period. Residual – difference between the raw data and the seasonal trend.

Fatal myocardial infarction rates declined throughout the study period (Figure 4). After accounting for seasonal effects, there was no demonstrable change in myocardial infarction mortality rates following the onset of the COVID-19 pandemic.

The pattern of fatal stroke rates was similar to that of all-cause mortality, with rates declining during 2018 to mid-2019, followed by an increase in the second half of 2019, which continued throughout 2020 (Figure 4). The peak in stroke mortality in August 2020 (23 deaths per 10,000 person-months) was the same as that of September 2017 (24 per 10,000 person-months). There were two peaks in stroke mortality during the COVID-19 pandemic, which coincided with peaks in all-cause mortality. During these peaks, a smaller proportion of strokes were recorded in the primary position of the death certificate, revealing a greater shift towards stroke as a contributing, rather than the underlying cause of death (Supplementary Material, Appendix F).

### Causes of death during the COVID-19 pandemic

Between 1^st^ January 2020 and 1^st^ August 2021, 7,508/20,042 (37%) of participants died. The most commonly recorded underlying cause of death was dementia. COVID-19 was the second commonest underlying cause of death during this period (Figure 5). From the 7,508 deceased participants, 1,451 (19%) had COVID-19 listed in any position on the death certificate, and 1,286 (17%) had COVID-19 recorded as the underlying cause of death. The median age at death from COVID-19 during this period was the same as for death from any cause (87 years, IQR 81-92 years).

**Figure 5.**
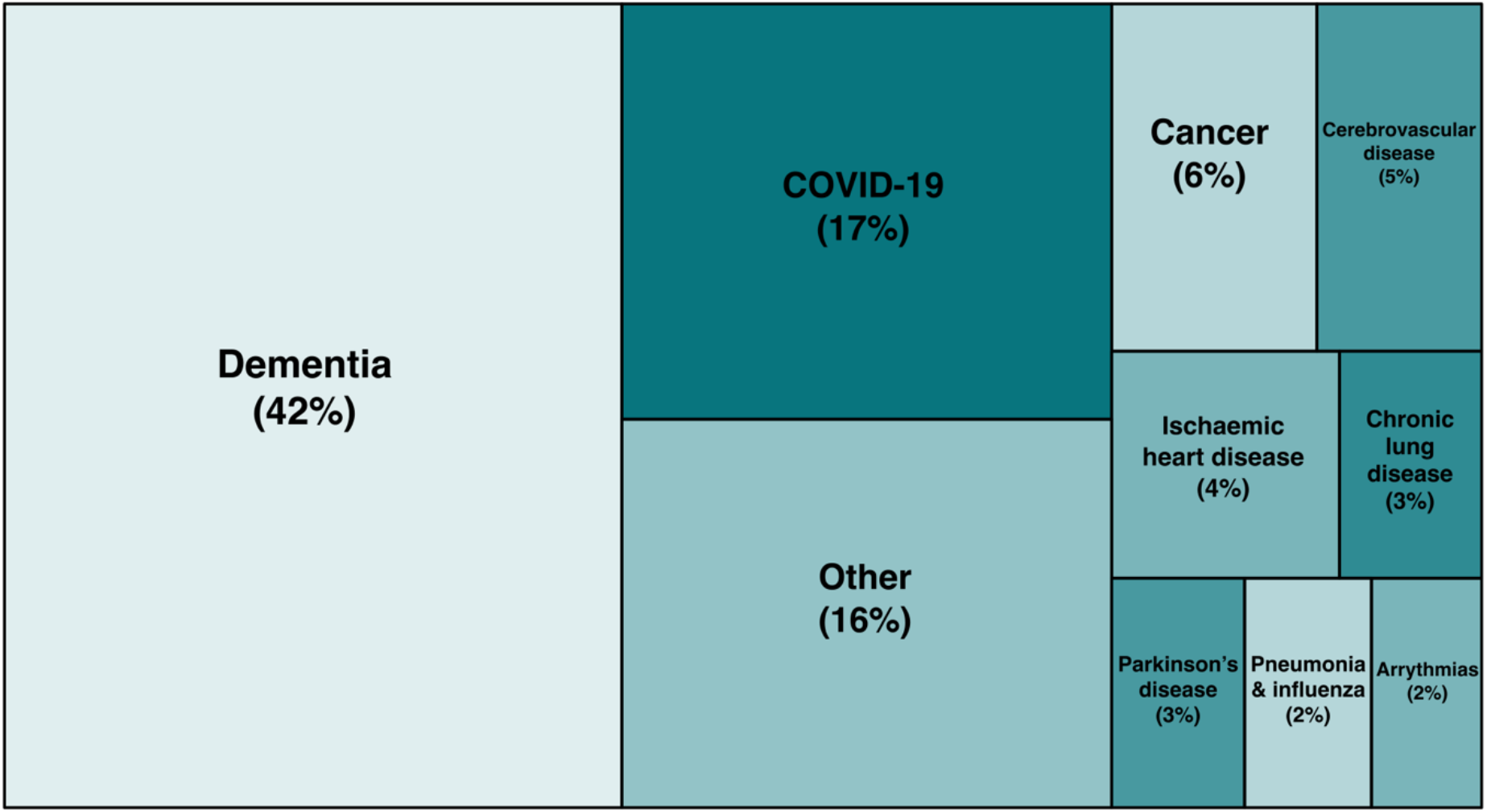
Treemap of leading underlying causes of death in people with dementia from January 2020 onwards.

## DISCUSSION

We used routinely-collected health data to examine trends in antipsychotic prescribing and mortality before and during the COVID-19 pandemic. Antipsychotic prescribing rates for people with dementia increased in 2019 and continued to increase during 2020, but the absolute change was small and was on a background of lower rates of prescribing over the previous two years. Antipsychotic prescribing in people with a subtype diagnosis of Alzheimer’s disease increased throughout the five-year study period. From 2020 onwards, COVID-19 became the leading non-dementia cause of death in people with dementia.

### Antipsychotic prescribing before and during the COVID-19 pandemic

After accounting for seasonal effects on prescribing, we observed a small increase in the proportion of people with dementia prescribed antipsychotic medications following the onset of the COVID-19 pandemic. However, the increased rates of prescribing began in the months prior to the arrival of COVID-19, suggesting that the pandemic was not the only potential cause in the change of practice. The age-stratified analysis revealed a notable increase in antipsychotic prescribing in the youngest age group (60-64 years) during the five-year study period: the reasons for this finding are unclear, and merit further exploration.

In the subset of people with Alzheimer’s disease, antipsychotic prescribing rates increased steadily from 2016 onwards with a sharp increase in the latter half of 2019, which continued during 2020.

A recent report highlighted an increased proportion of dementia patients being prescribed antipsychotics in 2020 compared to previous years, based on publicly-available NHS safety data in England.^15^ The authors noted that the proportion of people with dementia prescribed an antipsychotic was 9·7% in March 2020, compared to 4·3% the year before. In contrast, our study identified that 12·9% of people with dementia were prescribed an antipsychotic drug in March 2020, compared to 12·5% the year before. Our study was based in Wales and the publicly available NHS data were from England. Although healthcare services are devolved to each UK nation, Wales and England use the same clinical guidance and standards, meaning it is unlikely that the variations in prescribing between the studies represent true differences between healthcare practices.^13^ Instead, it is likely that the higher proportion of people prescribed an antipsychotic in Wales represents methodological differences between the two studies – our study used a validated code list to identify dementia patients from primary care and hospital admissions data, whereas the NHS safety data solely uses primary care dementia registers, which are maintained by individual general practices. Although our study identified an increase in antipsychotic prescribing between 2019 and 2020, the use of a time series analysis revealed that, once seasonal variations were accounted for, the absolute increase was minimal, and occurred following particularly low prescribing rates over the previous two years.

Harrison et al. (2021) used US routinely-collected data to compare the proportion of people with dementia prescribed an antipsychotic in the 30 days prior to attendance at a healthcare organisation (e.g. hospital or primary care centre) in 2020, compared to 2019.^16^ In this study, 16·4% of patients were prescribed an antipsychotic during 2020, compared to 14·7% in 2019. The method of data collection in this setting only allowed for measuring antipsychotic use prior to a healthcare attendance, so the study could not assess rates of antipsychotic prescribing for people who did not attend healthcare centres. It is likely that, during 2020, people attending healthcare centres were more unwell on average compared to the previous year, given the concerns about nosocomial COVID-19 transmission. This may have had biased the findings towards increased rates of antipsychotic use in this study.

Sizoo et al. (2022) investigated antipsychotic prescribing rates in psychogeriatric residents with dementia in the Netherlands, before and during national lockdowns.^26^ The proportion of people prescribed an antipsychotic was high (21·0-22·9%), reflecting the known higher rates of antipsychotic use in care home populations. They did not identify a change in prescribing practices over the study period.

### COVID-19 Mortality

The analysis revealed a marked increase in all-cause mortality from late 2019 onwards, with two peaks during 2020 and early 2021, which coincided with the known peaks in COVID-19 transmission in the UK.^27^ Previous studies have demonstrated that people with dementia are at an increased risk of severe disease or death from COVID-19.^2,28^

From January 2020 to August 2021, COVID-19 became the leading non-dementia underlying cause of death in people with dementia. Based on our analysis, six percent of all people with dementia who were alive at the start of 2020 died of COVID-19 over the subsequent 19 months. This is likely to be a significant underestimate, due to undertesting of COVID-19 early in the pandemic in the UK, particularly for those not admitted to hospital, which likely led to underreporting of COVID-19 on death certificates.^29^

### Cardiovascular and cerebrovascular diseases

Stroke mortality trends largely followed that of all-cause mortality. Following a reduction in stroke mortality during 2018, we observed an increase in the second half of 2019, which continued throughout 2020, albeit with a lower peak than in 2017. During the peaks in mortality, we identified a shift towards stroke appearing as a contributing rather than the underlying cause of death. This may in part be due to the increased risk of stroke following COVID-19 infection.^30^ In contrast to stroke, we observed a reduction in the rate of myocardial infarction deaths during the study period, with no increase following the onset of COVID-19.

### Antipsychotic prescribing and risk of stroke

A recent meta-analysis identified that antipsychotic use in people with dementia was consistently associated with a 16% increased risk of stroke across cohort studies.^5^ Our study design could not determine whether there is a causal relationship between any changes in antipsychotic prescribing and resulting stroke and all-cause mortality outcomes. For example, antipsychotic medications can be prescribed to people who have delirium, which they develop because they are medically unwell. The disease causing the delirium may therefore lead to both an increased likelihood of antipsychotic use and increased mortality (confounding by indication). In addition, people may receive antipsychotics palliatively to ease distress (reverse causation). Similarly, although we suspect the majority of the observed increase in all-cause mortality is likely to have been caused by COVID-19 itself, we cannot prove this using this study design.

### Strengths and limitations

This study benefited from the application of a validated code list to multiple complementary datasets to identify people with dementia.^23^ We used individual-level, linked, near-nationwide routine health data to construct a population-based cohort. The use of time series analyses allowed us to account for seasonal variations in antipsychotic prescribing and mortality, providing a more nuanced interpretation of changes over the five-year period.

There are several limitations. We used mortality data to study fatal stroke and myocardial infarction as outcomes, to avoid issues with determining whether repeated codes reflect recurrent events or duplicate coding of previous events. This approach will significantly underestimate the true rates of stroke and myocardial infarction. Routinely-collected health data will not identify disease outcomes with perfect accuracy, and death certificate data often has a lower positive predictive value and sensitivity compared to other datasets.^22^ Given the lack of access to COVID-19 tests early in the pandemic in the UK, it is likely that the COVID-19 mortality results are underestimates.^29^

## Conclusion

There was a large increase in stroke and all-cause mortality in people with dementia during the COVID-19 pandemic. COVID-19 is now the commonest non-dementia cause of death in this population. There was a small absolute increase in antipsychotic prescribing in people with dementia during the pandemic, but this rise had begun beforehand. Prescribing rates in the youngest age group and in people with Alzheimer’s disease have increased steadily over the last five years. Deprescribing antipsychotic medications where possible remains an essential aspect of dementia care.

## Data Availability

The data used in this study are available in the SAIL Databank at Swansea University, Swansea, UK, but as restrictions apply they are not publicly available. All proposals to use SAIL data are subject to review by an independent Information Governance Review Panel (IGRP). Before any data can be accessed, approval must be given by the IGRP. The IGRP gives careful consideration to each project to ensure proper and appropriate use of SAIL data. When access has been granted, it is gained through a privacy protecting safe haven and remote access system referred to as the SAIL Gateway.

## Acknowledgements

This work is carried out with the support of the BHF Data Science Centre led by HDR UK (BHF Grant no. SP/19/3/34678). This study makes use of de-identified data held in the SAIL Databank and made available via the BHF Data Science Centre’s CVD-COVID-UK/COVID-IMPACT consortium. This work uses data provided by patients and collected by the NHS as part of their care and support. We would also like to acknowledge all data providers who make health relevant data available for research.

This study makes use of anonymised data held in the Secure Anonymised Information Linkage (SAIL) Databank. This work uses data provided by patients and collected by the NHS as part of their care and support. We would also like to acknowledge all data providers who make anonymised data available for research. We wish to acknowledge the collaborative partnership that enabled acquisition and access to the de-identified data, which led to this output. The collaboration was led by the Swansea University Health Data Research UK team under the direction of the Welsh Government Technical Advisory Cell (TAC) and includes the following groups and organisations: the SAIL Databank, Administrative Data Research (ADR) Wales, Digital Health and Care Wales (DHCW), Public Health Wales, NHS Shared Services Partnership (NWSSP) and the Welsh Ambulance Service Trust (WAST). All research conducted has been completed under the permission and approval of the SAIL independent Information Governance Review Panel (IGRP) project number 0911.

For the purpose of open access, the author has applied a CC-BY public copyright licence to any Author Accepted Manuscript version arising from this submission.

## Data availability

The North East -Newcastle and North Tyneside 2 research ethics committee provided ethical approval for the CVD-COVID-UK/COVID-IMPACT research programme (REC No 20/NE/0161) to access, within secure trusted research environments, unconsented, whole-population, de-identified data from electronic health records collected as part of patients’ routine healthcare.

The CVD-COVID-UK/COVID-IMPACT Approvals & Oversight Board (https://www.hdruk.ac.uk/projects/cvd-covid-uk-project/) granted approval to this project to access the data within the Secure Anonymised Information Linkage (SAIL) Databank. The de-identified data used in this study were made available to accredited researchers only. Those wishing to gain access to the data should contact in the first instance.

The data used in this study are available in the SAIL Databank at Swansea University, Swansea, UK, but as restrictions apply they are not publicly available. All proposals to use SAIL data are subject to review by an independent Information Governance Review Panel (IGRP). Before any data can be accessed, approval must be given by the IGRP. The IGRP gives careful consideration to each project to ensure proper and appropriate use of SAIL data. When access has been granted, it is gained through a privacy protecting safe haven and remote access system referred to as the SAIL Gateway. SAIL has established an application process to be followed by anyone who would like to access data via SAIL at https://www.saildatabank.com/application-process.

## SUPPLEMENTARY MATERIAL

Appendix A: Codes used to identify dementia cases

Appendix B: Drug codes

Appendix C: Codes used to identify ischaemic stroke and myocardial infarction

Appendix D: Prescribing rates of individual antipsychotic drugs and benzodiazepines

Appendix E: Antipsychotic prescribing rates, stratified by age group

Appendix F: Stroke and myocardial infarction mortality rates, stratified by coding position

