## Supplementary Material for "Antipsychotic prescribing and mortality in people with dementia before and during the COVID-19 pandemic: retrospective cohort study"

**Contents**

**Page**

**Appendix A: Codes used to identify dementia cases ………………………………………………………… 2**

**Appendix B: Drug codes ………………………………………………………………………………………. 5**

**Appendix C: Codes used to identify ischaemic stroke and myocardial infarction ………………………... 6**

**Appendix D: Prescribing rates of individual antipsychotic drugs and benzodiazepines ………………….. 8**

**Appendix E: Antipsychotic prescribing rates, stratified by age group ……………………………………. 10**

**Appendix F: Stroke and myocardial infarction mortality rates, stratified by coding position ………….. 11**

**Appendix A: Codes used to identify dementia cases**

**ICD-10**

| **Description** | **Code** | **Subtype** |
| --- | --- | --- |
| Dementia in Alzheimer's disease | F00 | AD |
| Dementia in Alzheimer's disease with early onset | F00.0 | AD |
| Dementia in Alzheimer's disease with late onset | F00.1 | AD |
| Dementia in Alzheimer's disease, atypical or mixed type | F00.2 | AD |
| Dementia in Alzheimer's disease, unspecified | F00.9 | AD |
| Alzheimer’s disease | G30 | AD |
| Alzheimer’s disease with early onset | G30.0 | AD |
| Alzheimer’s disease with late onset | G30.1 | AD |
| Other Alzheimer's disease | G30.8 | AD |
| Alzheimer's disease unspecified | G30.9 | AD |
| Vascular dementia | F01 | VD |
| Vascular dementia of acute onset | F01.0 | VD |
| Multi-infarct dementia | F01.1 | VD |
| Subcortical vascular dementia | F01.2 | VD |
| Mixed cortical and sub-cortical vascular dementia | F01.3 | VD |
| Other vascular dementia | F01.8 | VD |
| Vascular dementia, unspecified | F01.9 | VD |
| Binswanger's disease | I67.3 | VD |
| Dementia in Picks disease | F02.0 | FTD |
| Circumscribed brain atrophy | G31.0 | FTD |
| Sporadic Creutzfeldt-Jakob disease | A81.0 | O |
| Dementia in Creutzfeldt-Jacob disease | F02.1 | O |
| Dementia in Huntington’s disease | F02.2 | O |
| Dementia in Parkinson’s disease | F02.3 | O |
| Dementia in HIV disease | F02.4 | O |
| Mental and behavioural disorders due to use of alcohol - amnesic syndrome | F10.6 | O |
| Dementia in other diseases classified elsewhere | F02 | N |
| Dementia in other specified diseases classified elsewhere | F02.8 | N |
| Unspecified dementia | F03 | N |
| Delirium superimposed on dementia | F05.1 | N |
| Senile degeneration of brain | G31.1 | N |
| Other specified degenerative diseases of nervous system | G31.8 | N |

**Read version 2**

| **Description** | **Code** | **Subtype** |
| --- | --- | --- |
| [X] Dementia in Alzheimer's disease | Eu00. | AD |
| [X]Dementia in Alzheimer's disease with early onset | Eu000 | AD |
| [X]Dementia in Alzheimer's disease with late onset | Eu001 | AD |
| [X]Dementia in Alzheimer's disease, atypical or mixed type | Eu002 | AD |
| [X]Dementia in Alzheimer's disease, unspecified | Eu00z | AD |
| Alzheimer’s disease | F110. | AD |
| Alzheimer’s disease with early onset | F1100 | AD |
| Alzheimer’s disease with late onset | F1101 | AD |
| Senile degeneration of brain | F112. | AD |
| [X] Other Alzheimer's disease | Fyu30 | AD |
| Multi-infarct dementia | E004. | VD |
| Uncomplicated arteriosclerotic dementia | E0040 | VD |
| Arteriosclerotic dementia with delirium | E0041 | VD |
| Arteriosclerotic dementia with paranoia | E0042 | VD |
| Arteriosclerotic dementia with depression | E0043 | VD |
| Arteriosclerotic dementia NOS | E004z | VD |
| [X]Vascular dementia | Eu01. | VD |
| [X]Vascular dementia of acute onset | Eu010 | VD |
| [X]Multi-infarct dementia | Eu011 | VD |
| [X]Other vascular dementia | Eu01y | VD |
| [X]Vascular dementia, unspecified | Eu01z | VD |
| Cerebral degeneration due to cerebrovascular disease | F11x2 | VD |
| Binswanger's disease | F21y2 | VD |
| [X] Lewy body dementia | Eu025 | DLB |
| Lewy body disease | F116. | DLB |
| [X] Dementia in Picks disease | Eu020 | FTD |
| Pick's disease | F111. | FTD |
| Frontotemporal degeneration | F118. | FTD |
| Jakob-Creutzfeldt disease | A411. | O |
| Sporadic Creutzfeldt-Jakob disease | A4110 | O |
| Alcoholic dementia, NOS | E012. | O |
| Dementia in conditions EC | E041. | O |
| [X] Dementia in other diseases classified elsewhere | Eu02. | O |
| [X] Dementia in Creutzfeldt-Jacob disease | Eu021 | O |
| [X] Dementia in Huntington’s disease | Eu022 | O |
| [X] Dementia in Parkinson’s disease | Eu023 | O |
| [X] Dementia in HIV disease | Eu024 | O |
| [X]Dementia in other specified diseases classified elsewhere | Eu02y | O |
| [X]Mental and behavioural disorders due to use of alcohol: amnesic syndrome | Eu106 | O |
| [X]Mental and behavioural disorders due to use of alcohol: residual and late-onset psychotic disorder | Eu107 | O |
| Cerebral degeneration due to Jacob-Creutzfeldt disease | F11x7 | O |
| Cerebral degeneration due to Parkinson’s disease | F11x9 | O |
| Corticobasal degeneration | F11y2 | O |
| H/O: dementia | 1461. | N |
| Assessment of psychotic and behavioural symptoms of dementia | 38C13 | N |
| GDS level 4 - moderate cognitive decline | 3AE3. | N |
| GDS level 5 - moderately severe cognitive decline | 3AE4. | N |
| GDS level 6 - severe cognitive decline | 3AE5. | N |
| GDS level 7 - very severe cognitive decline | 3AE6. | N |
| Dementia monitoring | 66h.. | N |
| Dementia annual review | 6AB.. | N |
| Dementia medication review | 8BM02 | N |
| Shared care – prescribing drug for dementia | 8BM50 | N |
| Shared care – prescribing drug for dementia declined | 8BM60 | N |
| Antipsyc drug therapy dementia | 8BPa. | N |
| Dementia advance care plan | 8CMe0 | N |
| Review of dementia advance care plan | 8CMG2 | N |
| Dementia care plan | 8CMZ. | N |
| Dementia care plan agreed | 8CMZ0 | N |
| Dementia care plan reviewed | 8CMZ1 | N |
| Dementia care plan declined | 8CMZ2 | N |
| Dementia care plan review declined | 8CMZ3 | N |
| Dementia advance care plan agreed | 8CSA. | N |
| Referral to dementia care advisor | 8Hla. | N |
| Dementia adv care plan declnd | 8IAe0 | N |
| Dementia advance care plan review declined | 8IAe2 | N |
| Exception reporting: dementia quality indicators | 9hD.. | N |
| Excepted from dementia quality indicators: patient unsuitable | 9hD0. | N |
| Excepted from dementia quality indicators: informed dissent | 9hD1. | N |
| Dementia monitoring administration | 9Ou.. | N |
| Dementia monitoring first letter | 9Ou1. | N |
| Dementia monitoring second letter | 9Ou2. | N |
| Dementia monitoring third letter | 9Ou3. | N |
| Dementia monitoring verbal invite | 9Ou4. | N |
| Dementia monitoring telephone invite | 9Ou5. | N |
| Senile and presenile organic psychotic condition | E00.. | N |
| Uncomplicated senile dementia | E000. | N |
| Pre-senile dementia | E001. | N |
| Uncomplicated pre-senile dementia | E0010 | N |
| Pre-senile dementia with delirium | E0011 | N |
| Pre-senile dementia with paranoia | E0012 | N |
| Pre-senile dementia with depression | E0013 | N |
| Pre-senile dementia NOS | E001z | N |
| Senile dementia with depressive or paranoid features | E002. | N |
| Senile dementia with paranoia | E0020 | N |
| Senile dementia with depression | E0021 | N |
| Senile dementia with depressive or paranoid features NOS | E002z | N |
| Senile dementia with delirium | E003. | N |
| Drug induced dementia | E02y1 | N |
| [X]Sub-cortical vascular dementia | Eu012 | N |
| [X]Mixed cortical and sub-cortical vascular dementia | Eu013 | N |
| [X] Unspecified dementia | Eu02z | N |
| [X] Delirium superimposed on dementia | Eu041 | N |

**Table A1. ICD-10 and Read version 2 codes used to identify dementia and its subtypes.**

AD – Alzheimer’s disease, VD – vascular dementia, FTD – frontotemporal dementia, DLB – Dementia with Lewy Bodies, O – other dementia subtype, N – no subtype specified.

For details of code list derivation and validation, see *Wilkinson, T. et al. Identifying dementia outcomes in UK Biobank: a validation study of primary care, hospital admissions and mortality data. Eur J Epidemiol* ***34****, 557–565 (2019).*

**Appendix B: Drug codes (Read V2)**

| **Code** | **Drug** | **Other indications** |
| --- | --- | --- |
| **Antipsychotics** | | |
| d4t.. | Amisulpride |  |
| d4v.. | Aripiprazole |  |
| d42.. | Benperidol |  |
| d41.. | Chlorpromazine |  |
| d4l.. | Clozapine |  |
| d45.. | Flupentixol |  |
| d46.. | Fluphenazine |  |
| d47.. | Haloperidol |  |
| d48.. | Levomepromazine | Antiemetic in palliative care |
| d4r.. | Olanzapine |  |
| d4w.. | Paliperidone |  |
| d4a.. | Pericyazine |  |
| d4b.. | Perphenazine |  |
| d4c.. | Pimozide |  |
| d4d.. | Prochlorperazine |  |
| d4e.. | Promazine |  |
| d4s.. | Quetiapine |  |
| d4p.. | Risperidone |  |
| d4f.. | Sulpiride |  |
| d4h.. | Trifluoperazine |  |
| d4n.. | Zuclopenthixol |  |
| **Benzodiazepines** | | |
| d22.. | Alprazolam |  |
| d23.. | Bromazepam |  |
| d2f.. | Buspirone hydrochloride |  |
| d24.. | Chlordiazepoxide | Alcohol withdrawal |
| d25.. | Chlormezanone |  |
| d26.. | Clobazam |  |
| dnc.. | Clobazam (epilepsy) |  |
| dn4.. | Clonazepam (epilepsy) |  |
| do2.. | Clonazepam (status epilepticus) |  |
| d27.. | Clorazepate dipotassium |  |
| o53.. | Diazepam (anaesthesia) |  |
| d21.. | Diazepam (anxiolytic) |  |
| do1.. | Diazepam (epilepsy) |  |
| d15.. | Flurazepam |  |
| d28.. | Hydroxyzine HCL |  |
| d16.. | Loprazolam |  |
| o56.. | Lorazepam (anaesthesia) |  |
| d2a.. | Lorazepam (anxiolytic) |  |

**Appendix C: Codes used to identify ischaemic stroke and myocardial infarction**

**Stroke – ICD-10**

| **Code** | **Description** |
| --- | --- |
| I63 | Cerebral infarction |
| I63.0 | Cerebral infarction due to thrombosis of precerebral arteries |
| I63.1 | Cerebral infarction due to embolism of precerebral arteries |
| I63.2 | Cerebral infarction due to unspecified occlusion or stenosis of precerebral arteries |
| I63.3 | Cerebral infarction due to thrombosis of cerebral arteries |
| I63.4 | Cerebral infarction due to embolism of cerebral arteries |
| I63.5 | Cerebral infarction due to unspecified occlusion or stenosis of cerebral arteries |
| I63.6 | Cerebral infarction due to cerebral venous thrombosis, nonpyogenic |
| I63.8 | Other cerebral infarction |
| I63.9 | Cerebral infarction, unspecified |

**Stroke – Read V2**

| **Code** | **Description** |
| --- | --- |
| G63y. | Other precerebral artery occlusion |
| G63y0 | Cerebral infarction due to thrombosis of precerebral arteries |
| G63y1 | Cerebral infarction due to embolism of precerebral arteries |
| G64.. | Cerebral arterial occlusion |
| G641. | Cerebral embolism |
| G640. | Cerebral thrombosis |
| G6400 | Cerebral infarction due to thrombosis of cerebral arteries |
| G6410 | Cerebral infarction due to embolism of cerebral arteries |
| G64z. | Cerebral infarction NOS |
| G64z0 | Brainstem infarction |
| G64z1 | Wallenberg syndrome |
| G64z2 | Left sided cerebral infarction |
| G64z3 | Right sided cerebral infarction |
| G64z4 | Infarction of basal ganglia |
| G6W.. | Cerebral infarction due to unspecified occl/stenosis of precerebral arteries |
| Gyu6G | Cerebral infarction due to unspecified occl/stenosis of precerebral arteries |
| G6X.. | Cerebral infarction due to unspecified occl/stenosis of cerebral arteries |
| Gyu63 | Cerebral infarction due to unspecified occl/stenosis of cerebral arteries |
| G6760 | Cerebral infarction due to cerebral venous thrombosis, non pyogenic |
| Gyu64 | Other cerebral infarction |

**Myocardial Infarction – ICD-10**

| **Code** | **Description** |
| --- | --- |
| I21 | Acute myocardial infarction |
| I21.0 | Acute transmural myocardial infarction of anterior wall |
| I21.1 | Acute transmural myocardial infarction of inferior wall |
| I21.2 | Acute transmural myocardial infarction of other sites |
| I21.3 | Acute transmural myocardial infarction of unspecified site |
| I21.4 | Acute subendocardial myocardial infarction |
| I21.9 | Acute myocardial infarction unspecified |
| I22 | Subsequent myocardial infarction |
| I22.0 | Subsequent myocardial infarction of anterior wall |
| I22.1 | Subsequent myocardial infarction of inferior wall |
| I22.8 | Subsequent myocardial infarction of other sites |
| I22.9 | Subsequent myocardial infarction of unspecified site |
| I23 | Certain current complications following acute myocardial infarction |
| I23.0 | Haemopericardium as current complication following acute myocardial infarction |
| I23.1 | Atrial septal defect as current complication following acute myocardial infarction |
| I23.2 | Ventricular septal defect as current complication following acute myocardial infarction |
| I23.3 | Rupture of cardiac wall without haemopericardium as current complication following acute myocardial infarction |
| I23.4 | Rupture of chordae tendineae as current complication following acute myocardial infarction |
| I23.5 | Rupture of papillary muscle as current complication following acute myocardial infarction |
| I23.6 | Thrombosis of atrium auricular appendage and ventricle as current complications following acute myocardial infarction |

**Myocardial infarction – Read V2**

| **Code** | **Description** |
| --- | --- |
| 323.. | ECG: myocardial infarction |
| 3233. | ECG: antero-septal infarct. |
| 3234. | ECG:posterior/inferior infarct |
| 3235. | ECG: subendocardial infarct |
| 3236. | ECG: lateral infarction |
| 323Z. | ECG: myocardial infarct NOS |
| 889A. | Diabetes mellitus insulin-glucose infusion in acute myocardial infarction |
| G30.. | Acute myocardial infarction |
| G300. | Acute anterolateral infarction |
| G301. | Other specified anterior myocardial infarction |
| G3010 | Acute anteroapical infarction |
| G3011 | Acute anteroseptal infarction |
| G301Z | Anterior myocardial infarction NOS |
| G302. | Acute inferolateral infarction |
| G303. | Acute inferoposterior infarction |
| G302. | Posterior myocardial infarction NOS |
| G305. | Lateral myocardial infarction NOS |
| G306. | True posterior myocardial infarction |
| G307. | Acute subendocardial infarction |
| G3070 | Acute non-Q wave infarction |
| G3071 | Acute non-ST segment elevation myocardial infarction |
| G308. | Inferior myocardial infarction NOS |
| G309. | Acute Q-wave infarct |
| G30B. | Acute posterolateral myocardial infarction |
| G30X. | Acute transmural myocardial infarction of unspecified site |
| G30X0 | Acute ST segment elevation myocardial infarction |
| G30y. | Other acute myocardial infarction |
| G30y0 | Acute atrial infarction |
| G30y1 | Acute papillary muscle infarction |
| G30y2 | Acute septal infarction |
| G30yz | Other acute myocardial infarction NOS |
| G30z. | Acute myocardial infarction NOS |
| G310. | Postmyocardial infarction syndrome |
| G31y1 | Microinfarction of heart |
| G35.. | Subsequent myocardial infarction |
| G350. | Subsequent myocardial infarction of anterior wall |
| G351. | Subsequent myocardial infarction of inferior wall |
| G353. | Subsequent myocardial infarction of other sites |
| G35X. | Subsequent myocardial infarction of unspecified site |
| G36.. | Certain current complications following acute myocardial infarction |
| G360. | Haemopericardium as current complication following acute myocardial infarction |
| G361. | Atrial septal defect as current complication following acute myocardial infarction |
| G362. | Ventricular septal defect as current complication following acute myocardial infarction |
| G363. | Rupture of cardiac wall without haemopericardium as current complication following acute myocardial infarction |

**Appendix D: Prescribing rates of individual antipsychotic drugs and benzodiazepines**

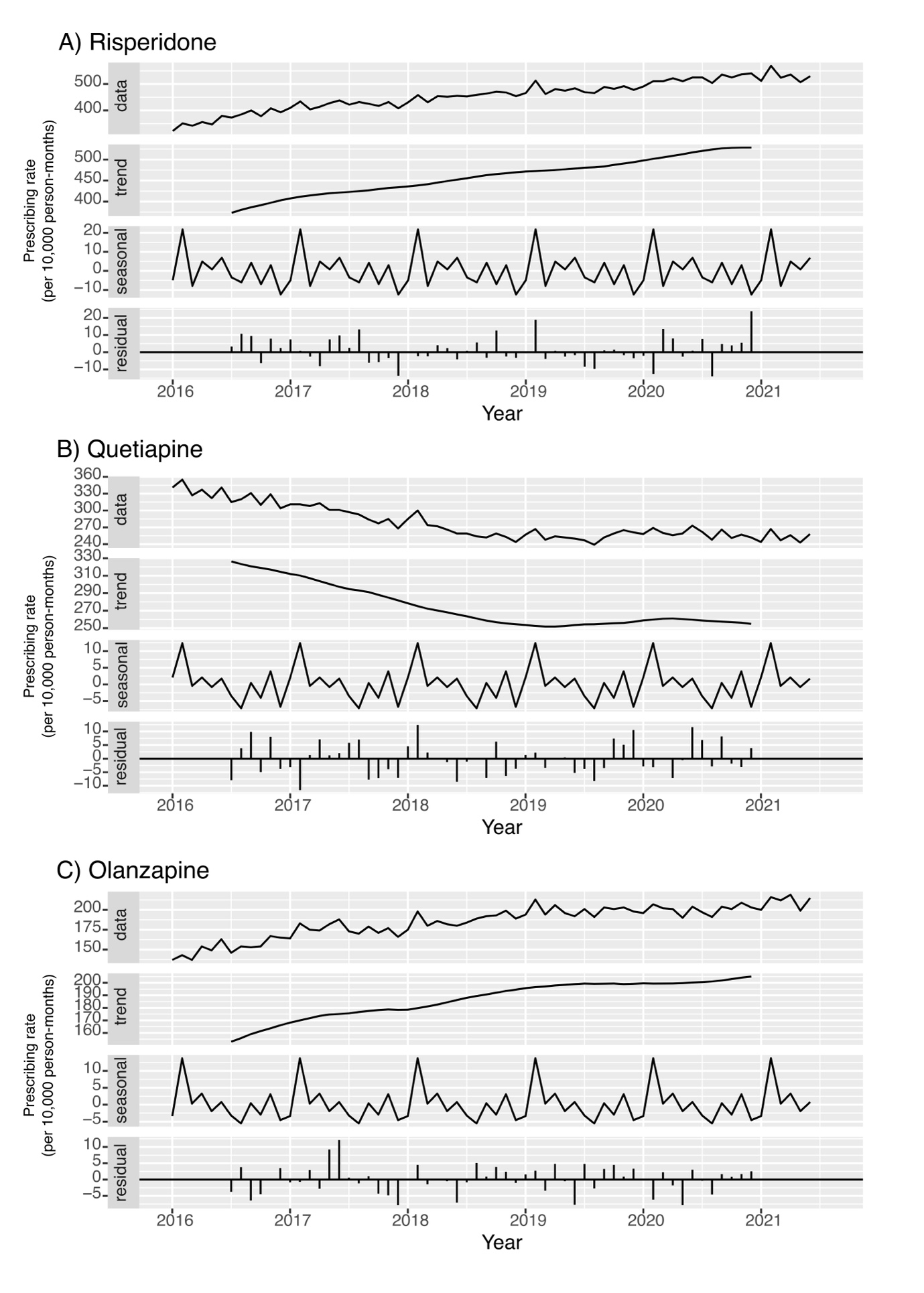

**Figure E.1 Time series analyses of prescribing rates for individual antipsychotic drugs**

Three most commonly prescribed antipsychotic drugs were risperidone (A), quetiapine (B) and olanzapine (C). Age-standardised, additive time series, accounting for seasonal trends in prescribing. Data – age-standardised prescribing rates. Trend – trend of prescribing rates, accounting for seasonal trends. Seasonal – seasonal trends in prescribing rates during the study period. Residual – difference between the raw data and the seasonal trend.

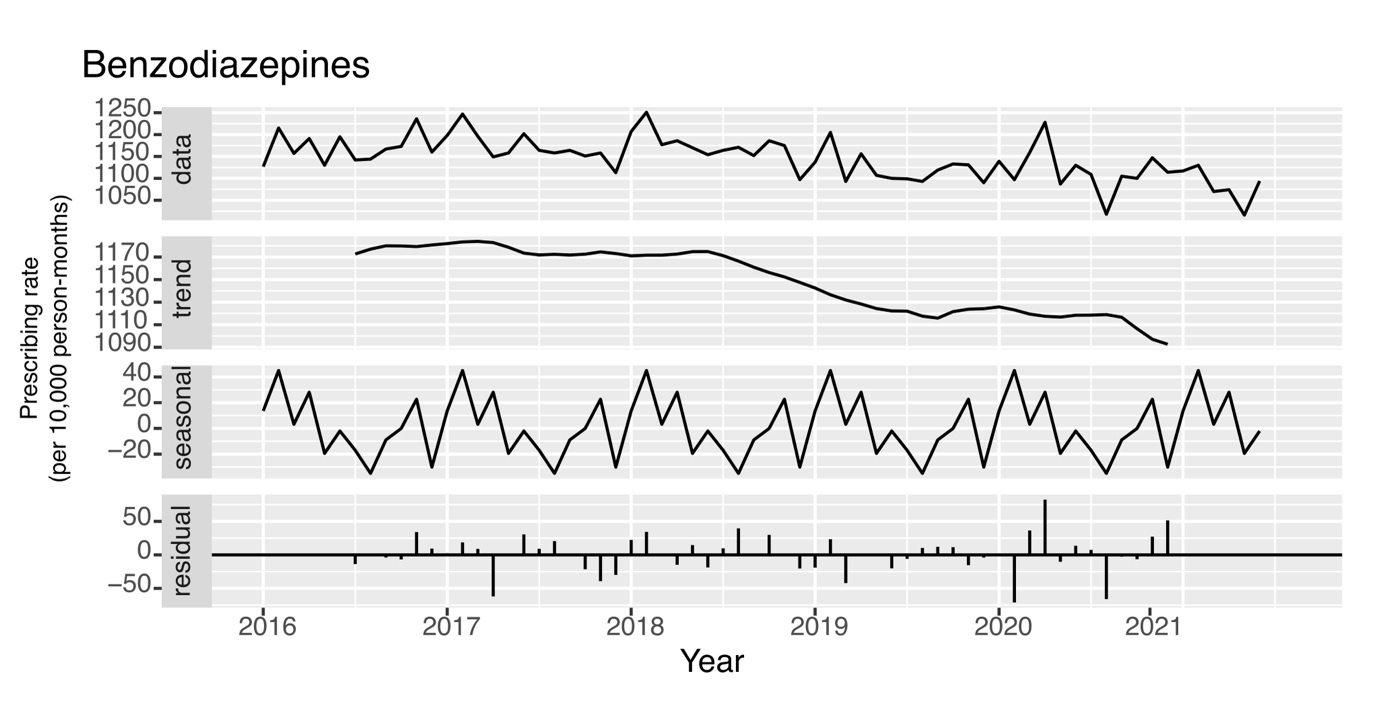

**Figure E.2 Time series analyses of prescribing rates for benzodiazepine drugs**

Benzodiazepine drugs displayed by way of comparison to antipsychotic prescribing practices throughout study period. Age-standardised, additive time series, accounting for seasonal trends in prescribing.

Data – age-standardised prescribing rates. Trend – trend of prescribing rates, accounting for seasonal trends. Seasonal – seasonal trends in prescribing rates during the study period. Residual – difference between the raw data and the seasonal trend.

**Appendix E: Antipsychotic prescribing rates, stratified by age group**

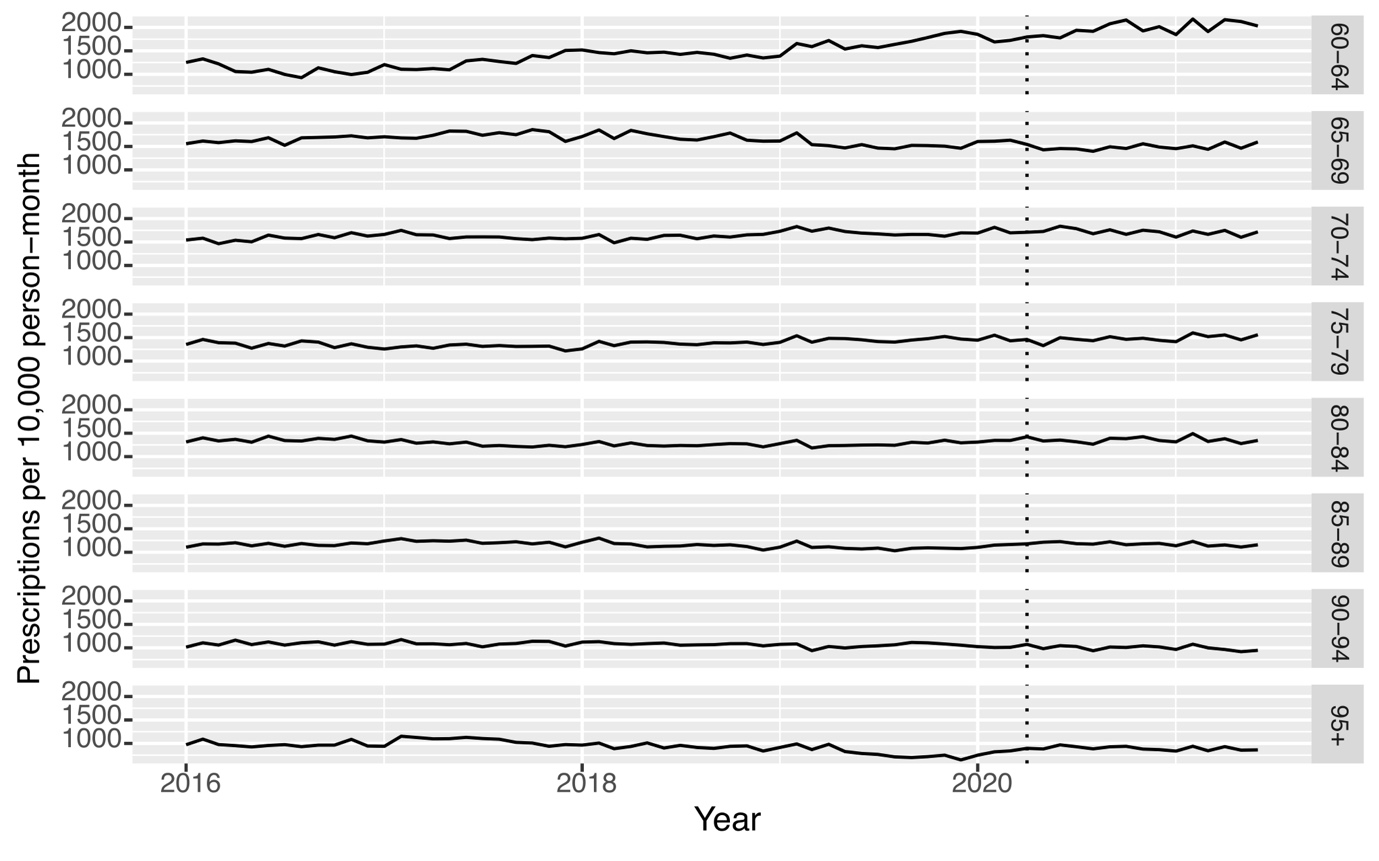

**Figure D.1. Age-standardised antipsychotic prescribing rates, stratified by age group.**

Ages grouped into 5-year bands. Dotted vertical line indicates March 2020, the month of the first UK-wide lockdown to reduce the spread of COVID-19.

**Appendix F: Stroke and myocardial infarction mortality rates, stratified by coding position**

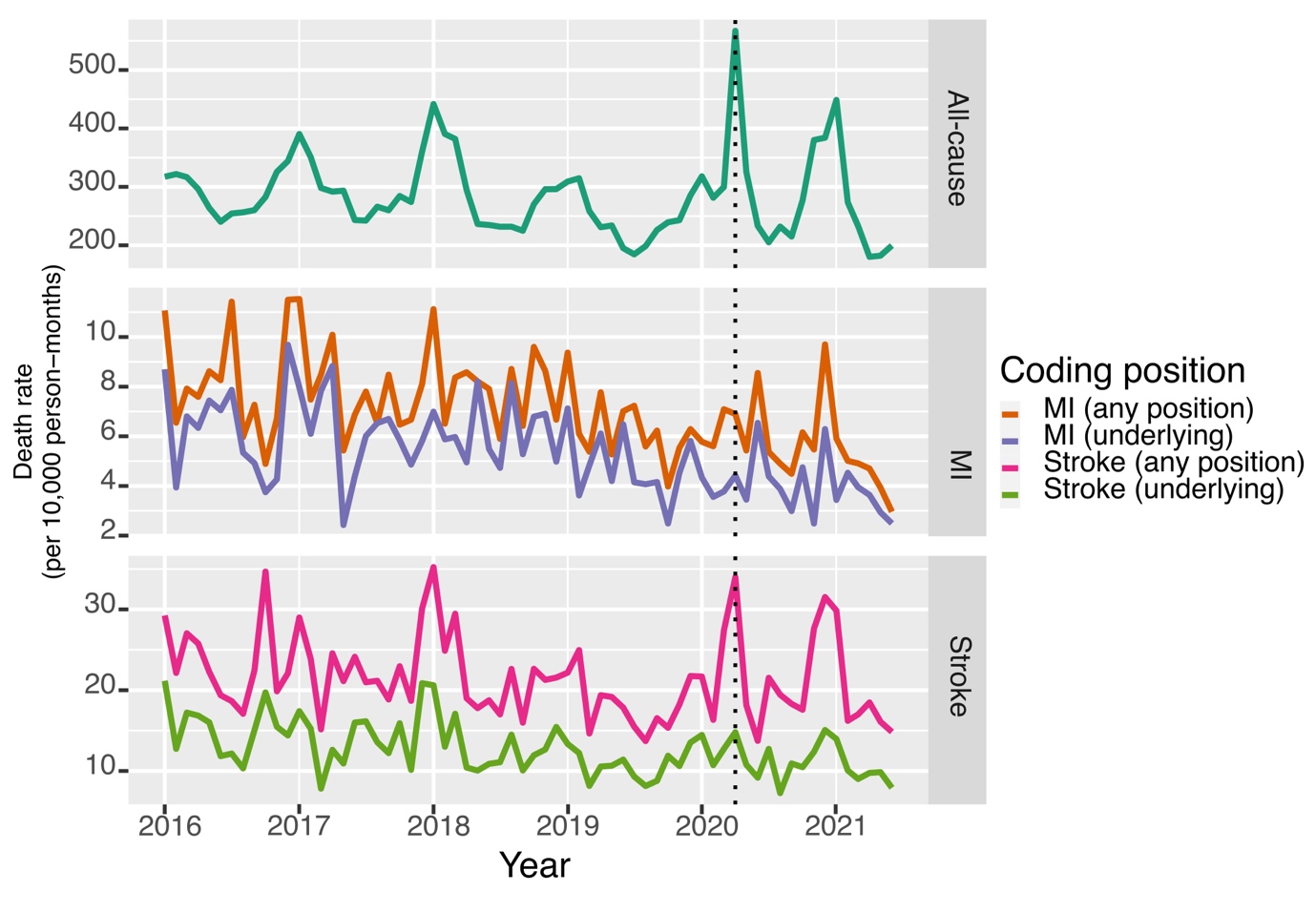

**Figure F1. Age-standardised all-cause, stroke and myocardial infarction mortality rates.**

For myocardial infarction and stroke, ‘underlying’ refers to the disease appearing in the primary position on the death certificate, whereas ‘any’ refers to the disease appearing anywhere on the death certificate (primary or secondary). Dotted vertical line indicates March 2020, the month of the first UK-wide lockdown to reduce the spread of COVID-19.
